# NutriRAG: Unleashing the Power of Large Language Models for Food Identification and Classification through Retrieval Methods

**DOI:** 10.1101/2025.03.19.25324268

**Authors:** Huixue Zhou, Lisa S. Chow, Lisa Harnack, Satchidananda Panda, Emily N.C. Manoogian, Minchen Li, Yongkang Xiao, Rui Zhang

**Author notes:** Corresponding author: Dr. Rui Zhang, PhD, Division of Computational Health Sciences, Department of Surgery, University of Minnesota, 11-132 Phillips-Wangensteen Building, 516 Delaware St SE, Minneapolis, MN 55455.

## Abstract

**Objective:** This study explores the use of advanced Natural Language Processing (NLP) techniques to enhance food classification and dietary analysis using raw text input from a diet tracking app.

**Materials and Methods:** The study was conducted in three stages: data collection, framework development, and application. Data were collected via the myCircadianClock app, where participants logged their meals in free-text format. Only de-identified food-related entries were used. We developed the NutriRAG framework, an NLP framework utilizing a Retrieval-Augmented Generation (RAG) approach to retrieve examples and incorporating large language models such as GPT-4 and Llama-2-70b. NutriRAG was designed to identify and classify user-recorded food items into predefined categories and analyzed dietary patterns from free-text entries in a 12-week randomized clinical trial (RCT: NCT04259632). The RCT compared three groups of obese participants: those following time-restricted eating (TRE, 8-hour eating window), caloric restriction (CR, 15% reduction), and unrestricted eating (UR).

**Results:** NutriRAG significantly enhanced classification accuracy and effectively identified nutritional content and analyzed dietary patterns, as noted by the retrieval-augmented GPT-4 model achieving a Micro F1 score of 82.24. Both interventions showed dietary alterations: CR participants ate fewer snacks and sugary foods, while TRE participants reduced nighttime eating.

**Conclusion:** By using AI, NutriRAG marks a substantial advancement in food classification and dietary analysis of nutritional assessments. The findings highlight NLP’s potential to personalize nutrition and manage diet-related health issues, suggesting further research to expand these models for wider use.

## INTRODUCTION

Nutrition plays a critical role in maintaining health and well-being. It plays an essential role for preventing and managing a multitude of chronic diseases, such as metabolic syndrome, obesity, cancer, hypertension, depression, and cardiovascular disease. [1–6]. Tracking food intake in the free-living setting, however, is a challenging task. The “gold-standard” for tracking food intake is the 24-hour dietary recall, however, these recalls represent “points in time” and do not necessary capture aggregate dietary patterns over time. A potential solution for capturing dietary intake is leveraging technology to capture food images and descriptions [7,8] and use various identification and classification techniques to analyze dietary habits.

The complexity of unstructured textual food descriptions poses significant challenges to classifying food intake. Recent advancements in Natural Language Processing (NLP), especially in semantic understanding and contextual analysis, offer promising solutions for automated food classification and nutritional assessment. NLP has been used to categorize food items and analyze packaged foods [16,17] and social media content [18]. For example, Hu et al. used the XGBoost algorithm to classify packaged foods into 24 distinct categories based on Health Canada’s Table of Reference Amounts [19]. Ma et al. developed a large dataset from the USDA Branded Food Products Database to classify various nutritional components like calories and proteins using a Multi-Layer Perceptron (MLP)-TF-SE approach [17].

There remains a knowledge gap in applying NLP to analyze personalized dietary patterns due to several limitations in existing databases and traditional classification methods. Databases such as the USDA Branded Food Products Database [20] and the University of Toronto Food Label Information and Price Database [21] predominantly focus on packaged items and offer limited coverage of fresh foods and recipes that are frequently part of dietary intakes.

Additionally, traditional NLP based classification, such as XGBoost and MLP are constrained due to their inability to capture complex patterns and dependencies in large-scale and high-dimensional data typical of nutritional studies, indicating a need for more sophisticated and comprehensive classification systems [22]. The emergence of generative large language models (LLMs), such as ChatGPT, which demonstrate improved performance on a variety of NLP tasks [23–30], presents new opportunities. These models have the potential to effectively classify textual descriptions of diverse food items.

In response to these challenges, our study introduces three significant contributions: 1) Advanced Food Classification System Development: We created a cutting-edge classification system that defines 51 unique food categories, capturing the complexity of contemporary dietary patterns and aligning with the latest trends in nutritional science. 2) Development and Application of the NutriRAG (Retrieval Augmented Generation-LLM for nutrition) Framework: We developed the NutriRAG framework, a few-shot, retrieval-based named entity recognition (NER) LLM framework tailored for food item identification and food class classification. This framework leverages technology-based mobile app records (from the myCircadianClock (mCC) smartphone app) to analyze personalized dietary patterns from text-based food entries. 3) Application and Testing of the NutriRAG Framework: We hypothesize that LLMs can effectively identify and categorize text-based food entries into predefined categories from a nutritional database. To validate this, we applied the NutriRAG framework to data from a randomized controlled trial with the intent to detect changes in dietary patterns over time.

## METHODS

As depicted in Figure 1, the study methodology is divided into three sections 1) Data Collection in the our previous study[31], where participants log their food intake using the mCC app [32], capturing the food textual descriptions; 2) Development of NutriRAG, which involves utilizing a Retrieval-Augmented Generation (RAG) strategy within a Large Language Model (LLM) to accurately process and identify food items and their classifications; and 3) Application of NutriRAG to analyze eating patterns (EO), which was defined as any food or drink consumption, including non-caloric beverages, separated by at least 15 minutes from other intake events[33]. This analysis helps to examine dietary habits and trends with the context of either a time-restricted eating or caloric restriction intervention.

**Figure 1.**
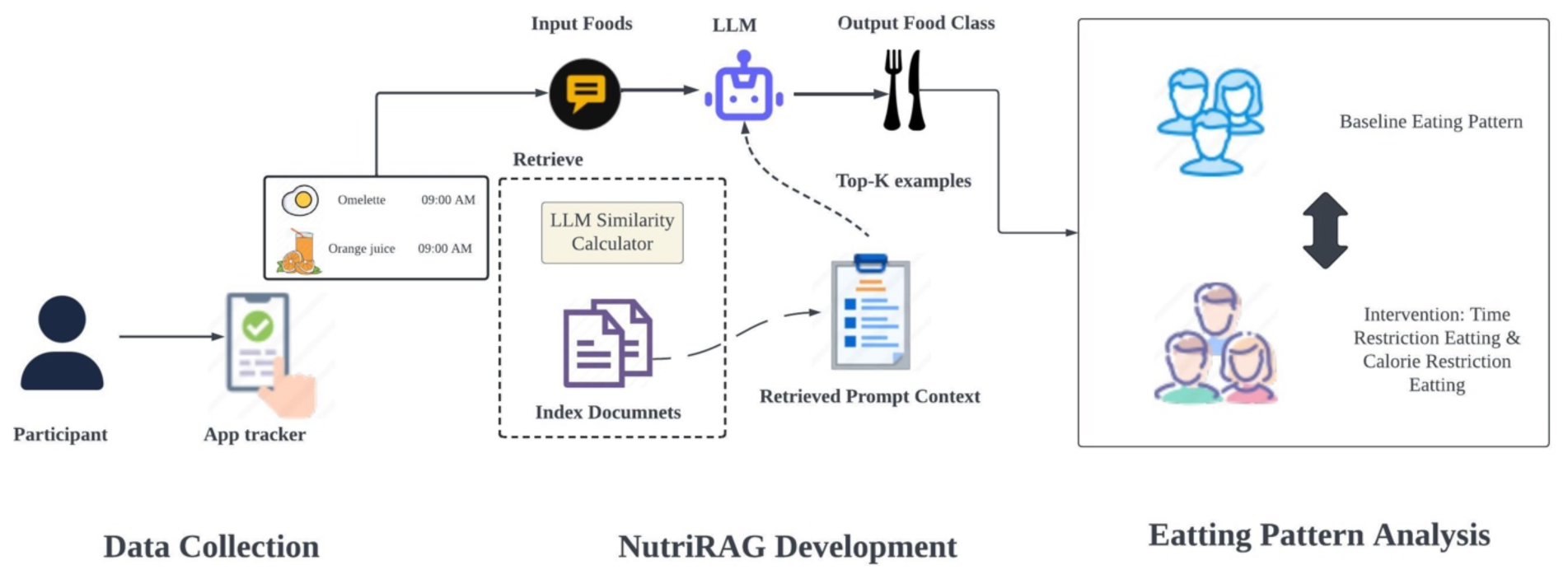
Overview of study.

### Design of Parent Study and Data Collection

This study builds upon a prior investigation [31] which is a randomized controlled trial (RCT), that compared three eating patterns in patients with obesity but without diabetes over 12 weeks: 8-hour Time-Restricted Eating (TRE), Caloric Restriction (CR, 15% daily reduction), and Unrestricted eating (UR). Participants from the Twin Cities metro area used the mCC mobile application to log their food and beverage intake, excluding water and medications [32]. Those with initial eating windows exceeding 12 hours qualified for the study and continued using the mCC app throughout the intervention period. Of 88 randomized participants, 81 completed the trial, with 77 providing analyzable data for this study: 27 in TRE, 25 in CR, and 25 in UR groups. The parent study received approval from the University of Minnesota’s Institutional Review Board (STUDY00008545) and the Salk Institutional Review Board (15-0003) for the mCC app. The trial was registered at ClinicalTrials.gov (NCT04259632), and all participants provided written informed consent.

For this specific study, we analyzed the data collected from the parent RCT, which largely came from the mcc app [32]. Participants logged their food consumption through images and free-text entries, receiving daily prompts to ensure consistent data input. All data, including timestamps, were encrypted, anonymized, and securely stored on a cloud server. For this analysis, we focused on the 32,825 free-text food entries recorded by 77 users.

The primary goal of this stage was to identify user recorded free text food items and classify each item into one of the predefined categories from the Nutrition Coordinating Center (NCC) Food Database, which contains 51 distinct food classes (Supplement Table 1). This NCC database provides a comprehensive classification system, meticulously detailing a wide array of food categories to encompass the diverse range of food items reported by users. To ensure our analysis was based on a robust and representative sample, we randomly selected 1,000 entries, which were manually categorized by NDSR certified staff against NCC database to serve as a “gold standard” benchmark. For instance, the input text “Carrots Short Ribs Potatoes” would be annotated to correctly identify each food item — “Carrots,” “Short Ribs,” “Potatoes”—with their respective NCC food classes as “Vegetables,” “Meat,” “Vegetables.” The dataset was then divided into subsets for training, validation, and testing, comprising 676, 168, and 182 entries, respectively.

### Development of NutriRAG

To improve the detection and sorting of nutritional data form the mCC app, we developed a new NLP framework NutriRAG, as depicted in Figure 2, which incorporates generative LLMs within a RAG strategy. This strategy leverages relevant documents during the generation process to enrich the model’s output contextually without requiring further tuning of model parameters. Our methodical approach is structured into four primary phases: 1) Query Formulation, where nutritional inquiries are constructed; 2) Retrieval and Prompt Context, where relevant documents are sourced; 3) LLM Processing, where the model processes the queries within the retrieved context; and 4) Organization of Nutritional Data Outputs, where we systematically arrange the LLM output data for analysis of dietary patterns.

**Figure 2.**
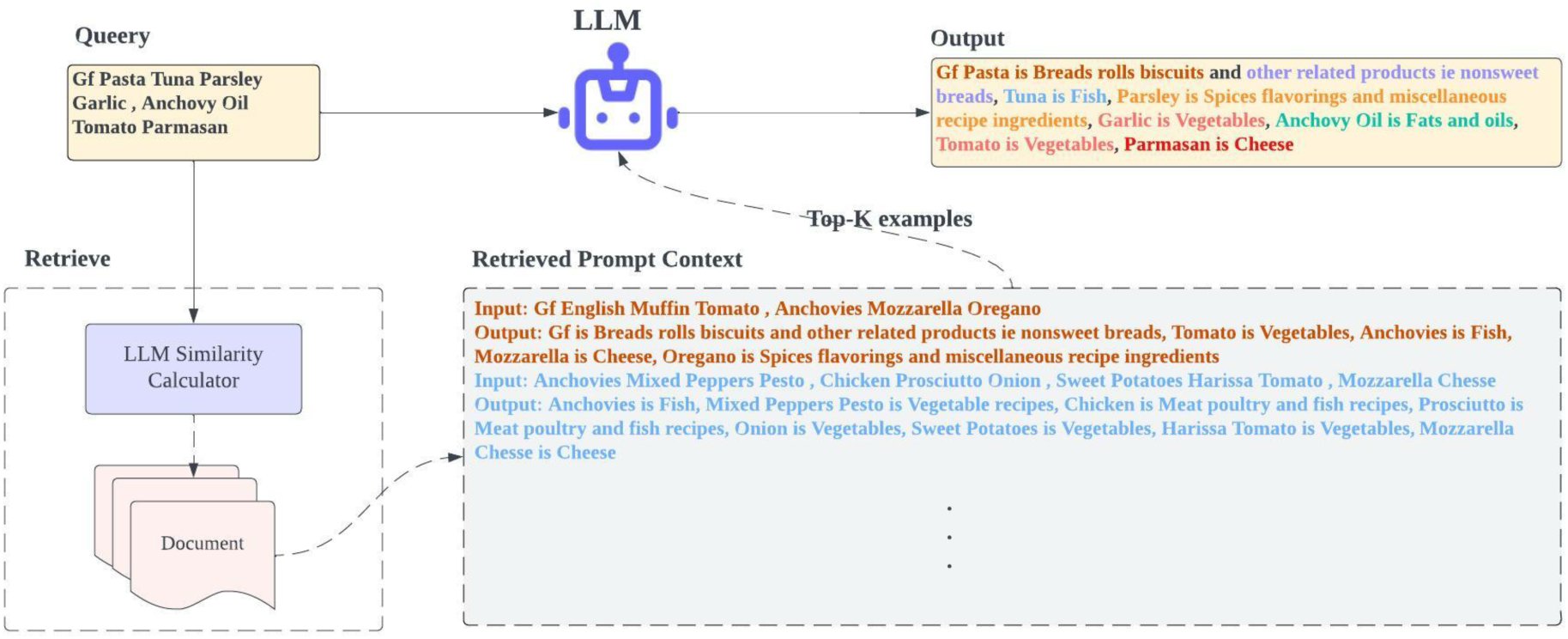
Framework of NutriRAG.

#### Query Formulation

As shown in Figure 2, The process begins with a “Query” phase, where a nutritional inquiry is composed, consisting of various food items and ingredients. For instance, a typical participant-entered text description might include “Gf Pasta Tuna Parsley Garlic Anchovy Oil Tomato Parmesan.”

#### Retrieval and Prompt Context

In the Retrieval and Prompt Context phase, we utilize an LLM Similarity Calculator to determine the score for example (*e_i_*) from valid dataset (*D*) for each query. This involves computing the cosine similarity score between the LLM embedding of the query text (*q_i_*) and *e_i_*, defined by the formula:

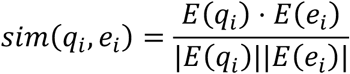

Here, *sim*(*q_i_*, *e_i_*) is the similarity score, and *E*(*q_i_*), *E*(*e_i_*) are the embeddings of the *q_i_* and *e_i_*, respectively. Next, we ranked the examples by their similarity scores and select the top k examples. This step ensures that the subsequent LLM processing is informed by contextually pertinent information, enhancing the accuracy of entity recognition. Once the examples are retrieved, the LLM is presented with a “Retrieved Prompt Context.” This context consists of a set of top-K examples illustrating input-output mappings of food items to their respective nutritional categories. These mappings are designed to act as a guiding framework for the LLM, providing clarity on the expected format and content of the output.

#### LLM Processing and Output Classification

The prompt of the LLM consists of instruction, example context and the query entry text. The instruction delineates the precise task to be executed by the model or specifies the desired response type. Meanwhile, the example context is sourced through *Retrieval and Prompt Context.* The LLM then processes prompt, mapping each component of the query entry to its appropriate nutritional category. This stage is crucial as it represents the core functionality of the NER system, wherein the LLM generates outputs such as “Tuna is Fish, Parsley is Spices flavorings and miscellaneous recipe ingredients.” The final LLM output was subsequently transformed into a structured format through string matching for each food class, thereby facilitating the integration of this information into databases or for further analytical processes.

#### Evaluation

For our evaluation, we employed advanced generative LLMs including GPT-3.5 and GPT-4, as well as open-source models such as Mixtral 8*7B [34] and Llama-2-70b [35], utilizing the RAG method. We tested retrieval from 1-20 examples and assessed three orders of retrieved examples: from highest to lowest similarity score, and in a random order. We compared these with base models selecting examples randomly and with fine-tuned pretrained language models such as BERT [36], PubmedBert [37], and BLUEBERT [38]. Performance was evaluated using micro precision, micro recall, and micro F1 score. The micro precision and micro recall are the measurement that considering the total number of true positives across all classes divided by the sum of true positives and false positives, and the total number of true positives across all classes divided by the sum of true positives and false negatives. The micro F1 is calculated by:

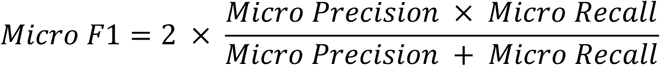

### Application of NutriRAG and Examination of Eating Patterns

Our study utilized the NutriRAG in the part I, to examine the eating patterns among the study participants and evaluated the intervention effect on the number of EO, which was defined as a distinct event when food or drink was documented ≥15 minutes apart from another EO [33]. Defined by our expert, food intake was classified into four daily segments: early morning (5 am - 10 am), midday (10:01 am - 3:00 pm), evening (3:01 pm - 9 pm), and late night (9:01 pm - 4:59 am), to evaluate the timing of dietary intake. The study analyzed the variance in dietary intake, especially with regards to late-night eating between baselines. During the 12-week intervention periods (UE, TRE, CR) of the RCT from the parent study, some EOs were tagged beverage, savory snacks, and high-added-sugar events using the LLM classifier from Step 1. Following the parent study, the study timeline was divided into three phases: baseline, post-randomization (from randomization to study completion), and at post-intervention (the last 2 weeks prior to study completion). The analysis focused on comparing the baseline with the post-randomization period and the baseline with the post-intervention period. Multiple linear regression analysis was used to evaluate the associations of and EOs with BMI and HOMA, after adjustments for age, sex. Results from the linear regression models are presented as standardized β-coefficients. In all analyses, the level of statistical significance was set at P < 0.05. Data were analyzed using R studio 4.1.2.

## RESULTS

### RCT results

As previously published [31], the TRE group maintained a 10.0-hour eating window (9.2 to 10.9) during the 12-week intervention. In the final two weeks of the intervention (Week 10-12), the TRE group’s eating window decreased to 9.1 hours (8.4 to 9.7), representing a 5.1-hour reduction (4.5 to 5.7) from baseline. Results from the 24-hour dietary recall data obtained at baseline and end-intervention showed average caloric reductions of 16.5% (−24.8 to −8.2) in the TRE group (N=28), 8.7% (−21.8 to 4.5) in the CR group (N=19), and 1.2% (−12.7 to 10.2) in the UE group (N=26). These reductions were not significantly different between groups.

This study analyzed available data from 77 participants (57% female) with a mean age of 43.3 ± 10.6 years. At baseline, participants averaged 105.6 ± 1.97 kg in weight, 173.0 ± 9.39 cm in height, and had a BMI of 36.4 ± 5.1 kg/m². Mean HOMA was 4.4 ± 3.88. Baseline daily eating occasions showed no significant differences between groups (P>0.05).

### NutriRAG

The results are described in Table 2. In the evaluation of various NER models, the baseline performances of Bert, BLUEBert, and PubmedBert were moderately effective, with Micro F1 scores of 59.38, 53.29, and 58.22, respectively. Notably, the innovative application of RAG significantly enhanced model performance across all configurations. For instance, while the standard GPT4 model achieved a Micro F1 of 73.84, its RAG augmented counterpart produced a substantially higher score of 82.24. For example, the standard GPT-4 model attained a Micro F1 score of 73.84, whereas its RAG-augmented version achieved a significantly higher score of 82.24. Likewise, retrieval-augmented versions of the Mixtral 8*7b models demonstrated notable improvements, reaching a Micro F1 score of 77.93—the highest among open-source models, surpassing the RAG-augmented GPT-3.5. Based on the model performance of Part I, we utilized the NutriRAG with Mixtral 8*7B, which achieved the best performance among the open-source models, to process the text-based food information for each participant.

**Table 1.** Performance of Models in Food Classification.

| Model | Micro precision | Micro Recall | Micro F1 |
| --- | --- | --- | --- |
| Bert | 56.36 | 62.74 | 59.38 |
| BLUEBert | 49.84 | 57.26 | 53.29 |
| PubmedBert | 56.82 | 59.71 | 58.22 |
| Random examples Llama-2-70b | 37.70 | 71.72 | 49.42 |
| Random examples Mixtral 8*7b | 60.87 | 75.58 | 67.43 |
| Random examples GPT3.5 | 64.54 | 66.43 | 65.47 |
| Random examples GPT4 | 75.97 | 71.53 | 73.84 |
| (UMN) Retrieval-Augmented Llama-2-70b | 68.81 | 58.75 | 63.38 |
| (UMN) Retrieval-Augmented Mixtral 8*7b | 76.07 | 79.89 | <u>77.93*</u> |
| (UMN) Retrieval-Augmented GPT3.5 | 75.00 | 80.29 | 77.55 |
| (UMN) Retrieval-Augmented GPT4 | 79.10 | 85.64 | **82.24\*** |

\* Note: Bold text indicates the highest values, while underscored text denotes the second.

**Table 2.**
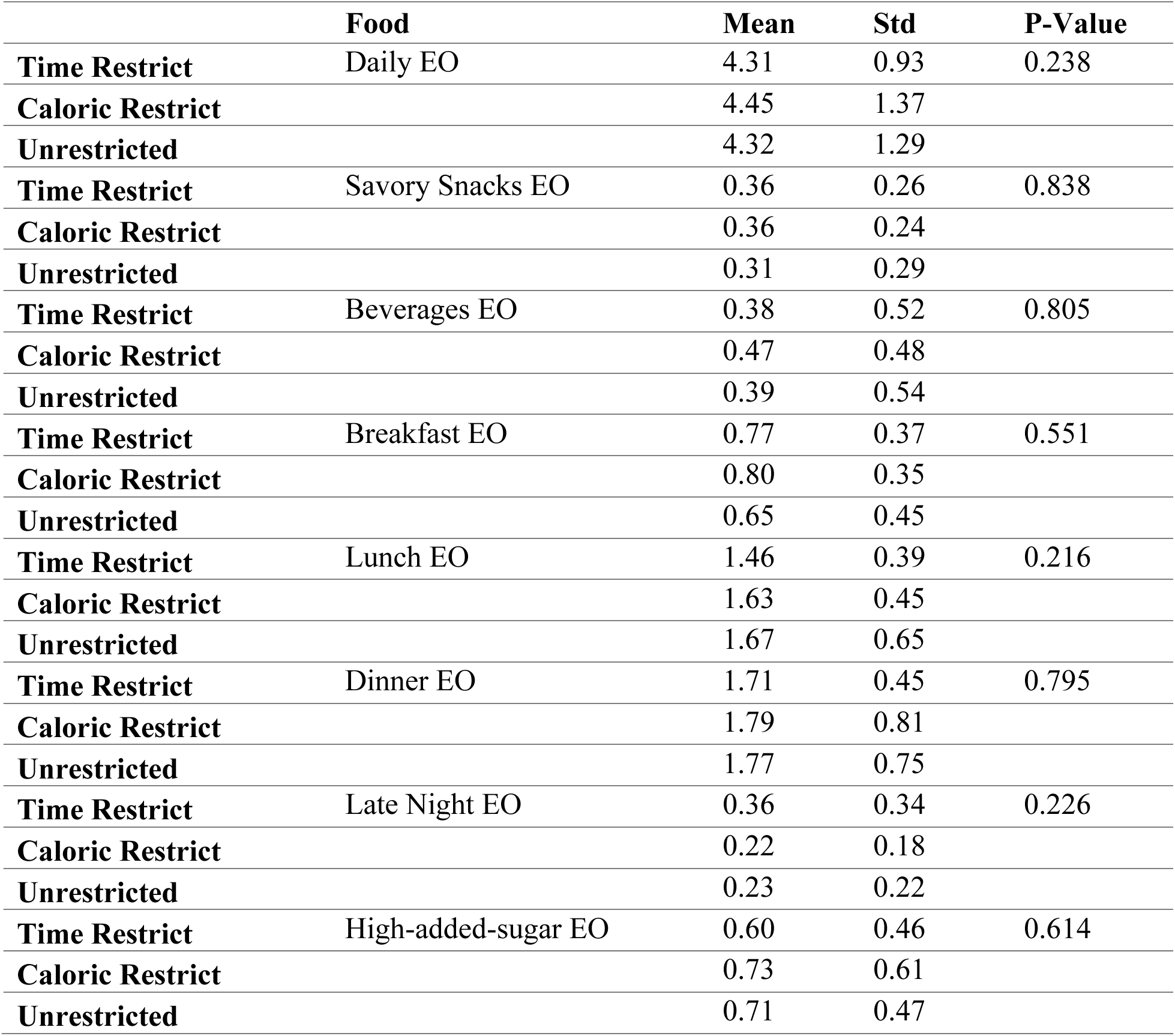
Baseline data.

### Examination of Eating Patterns

Post-randomization and post-intervention assessments revealed statistically significant differences in daily EOs for breakfast, lunch, late-night, and high-added-sugar events across the three groups (figure 3 and 4).

**Figure 3.**
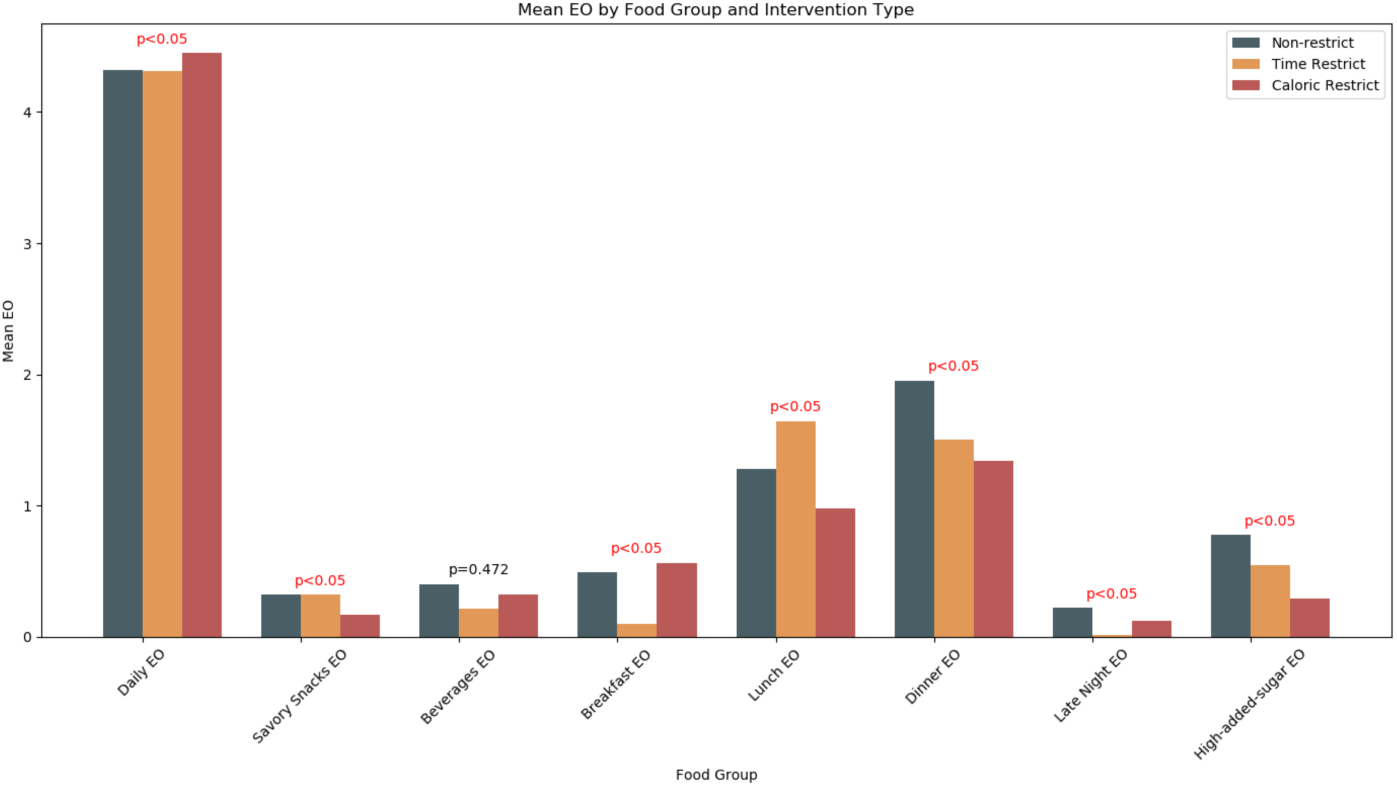
ANOVA of Different Group Post-randomization.

**Figure 4.**
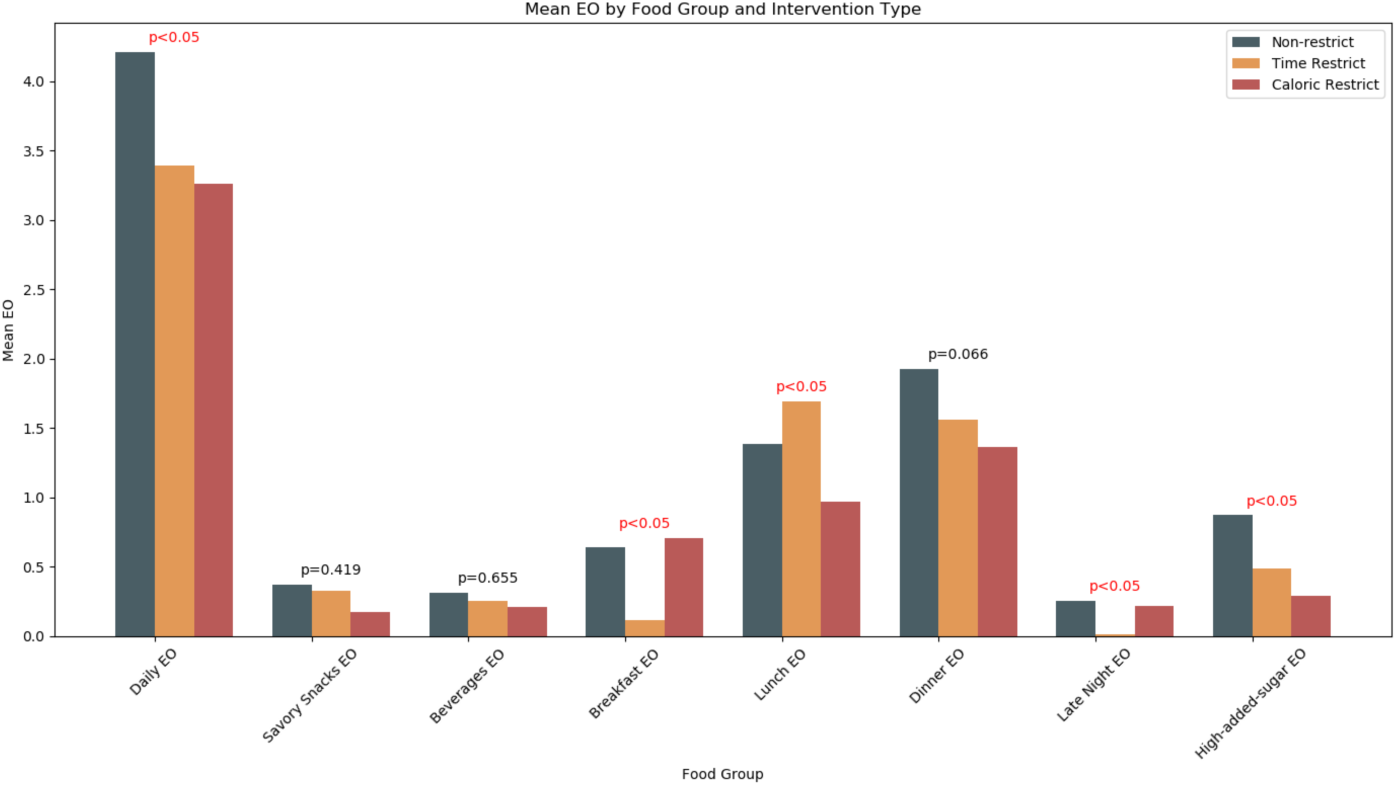
ANOVA of Different Group of Post-intervention.

For the UE group, the average daily EOs were 4.32 ±1.29 at baseline, 4.10 ±1.44 during the post-randomization, and 4.21 ±1.57 post-intervention. There were no significant differences in daily EOs when comparing baseline to post-randomization (P= 0.598) and baseline to post-intervention (P=0.887). Similarly, no significant changes were observed in the average number of daily EOs for savory snacks, beverages, late-night eating, and high-added-sugar foods between baseline and post-randomization (as shown in Figure 5) or between baseline and post-intervention (as shown in Figure 6).

**Figure 5.**
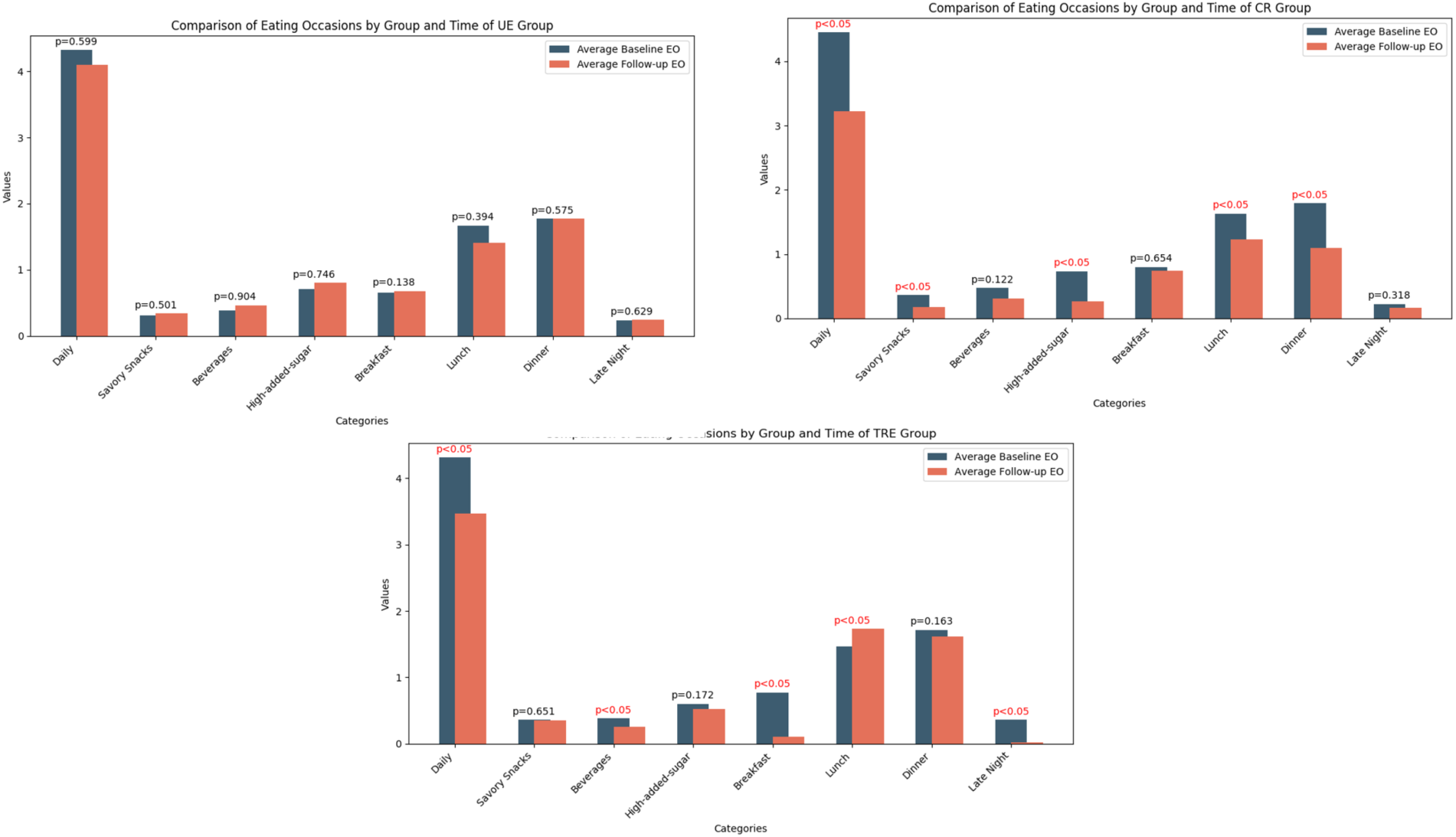
Paired T-Test of Baseline EOs and Post-randomization EOs.

**Figure 6.**
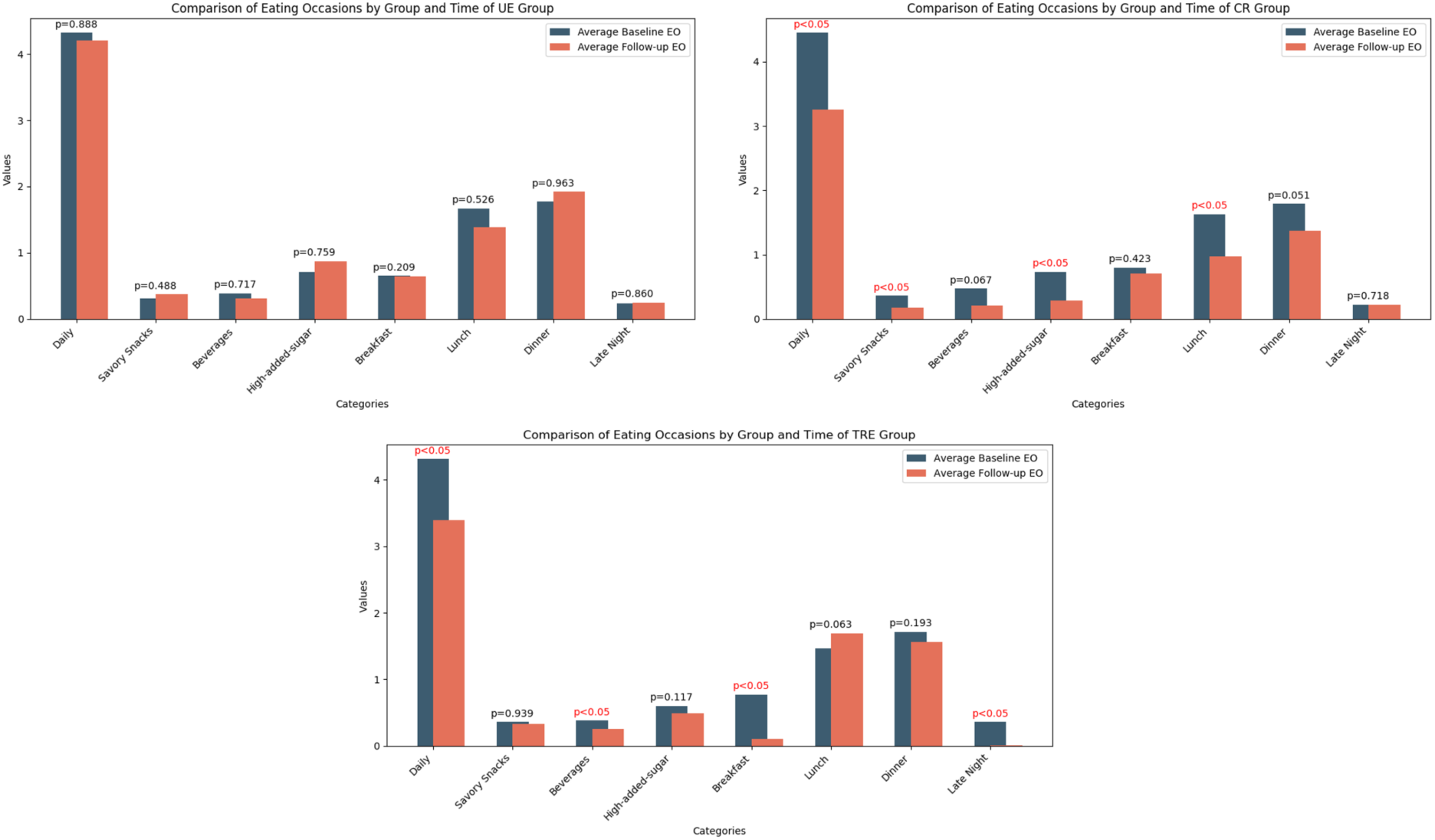
Paired T-Test of Baseline EOs and Post-intervention EOs.

Both the TRE and CR groups altered their dietary habits during the study. Initially, the average daily eating occasions (EOs) for TRE were 4.31 ±0.93, which decreased to 3.47 ±0.88 during the post-randomization and 3.39 ±1.03 at end-intervention. For the CRE group, the average daily EOs were 4.45 ±1.37 at baseline, reducing to 3.22 ±0.82 during the post-randomization and slightly rising to 3.26 ±0.97 at end-intervention. Significant reductions in daily EOs were observed in the TRE group from between baseline and during the post-randomization (P<0.001) and between baseline and end-intervention (P=0.001). Similar significant changes were noted in the CR group over these periods (P<0.001 for both comparisons).

The CR group decreased their consumption of savory snacks (P=0.003), and high-added-sugar foods (P<0.001). The TRE group similarly reduced their beverage intake (P=0.032) when during the post-randomization. In terms of meal timing, the CR group cut down on lunch (P=0.002) and dinner (P<0.001), while the TRE group lessened breakfast (P<0.001), lunch (P=0.011), and late-night eating (P<0.001).

These changes persisted into the end-intervention period. When comparing baseline to post-intervention, the CR group continued to consume fewer savory snacks (P=0.013), and high-added-sugar foods (P=0.003). The TRE group maintained reduced consumption of beverages (P=0.047), as illustrated in Figure 6. Regarding timing, the CR group still ate less at lunch (P=0.002), and the TR group continued to reduce breakfast and late-night eating (both P<0.001). In exploring the relationship between baseline metabolic parameters and eating occasions, we observed a link between baseline insulin resistance and dietary patterns. Notably, insulin sensitivity, characterized by higher HOMA values, inversely correlated with breakfast EO (P=0.019) and dinner EO (P=0.013). Specifically, for each one-unit increase in an EO at breakfast level, there was a decrease of 1.71 units in the HOMA value. This suggests that increased EO during the breakfast time period may be associated with lower insulin sensitivity. Conversely, no significant relationships were observed between eating occasions and BMI or lipid profiles.

**Table 3.** Linear Regression Analysis Results for HOMA.

| **Variable** | **Estimate** | **Std. Error** | **P-value** |
| --- | --- | --- | --- |
| **Age** | -0.002 | 0.025 | 0.938 |
| **Gender Male** | 0.492 | 0.548 | 0.372 |
| **BMI** | 0.193 | 0.051 | **<0.001** |
| **Breakfast EO** | -1.708 | 0.712 | **0.019** |
| **Lunch EO** | -0.466 | 0.636 | 0.466 |
| **Dinner EO** | -1.365 | 0.532 | **0.013** |
| **Late Night EO** | 0.769 | 1.063 | 0.472 |
| **High-added-sugar EO** | 0.666 | 0.639 | 0.301 |

## DISCUSSION

This study introduces NutriRAG, a novel approach integrating retrieval augmented generative LLM for the classification of food items from textual descriptions. Our findings demonstrate that NutriRAG significantly enhances the performance of standard LLMs in the domain of nutritional science.

The improved performance of retrieval-augmented models, as shown in our results, underscores the value of contextual enrichment in model training. For example, the Retrieval-Augmented GPT-4 model outperformed its standard counterparts by achieving a Micro F1 score of 82.24, compared to 73.84 for the non-augmented GPT-4. This enhancement can be attributed to the RAG method’s ability to dynamically incorporate relevant external data during the generation process, thus enabling the model to make more informed predictions based on a broader knowledge base.

The application of the mCC APP for tracking eating windows [31,39], complemented by the annotation of pre-processed entity texts by human nutrition experts [40], has underscored the integration of technological tools in dietary research. NutriRAG’s deployment in real-world scenarios further highlights its utility, as it a categorizes participant-entered free-text food items logged in the mCC app relative to an established nutritional database to calculate detailed nutritional profiles. Beyond its immediate application in organizing the free-text entries for the mCC app, NutriRAG’s capabilities extend to a broad array of technologies that handle free-text dietary input. This makes it an invaluable tool across various platforms where users log food intake, allowing for the transformation of raw textual entries into structured nutritional data. By categorizing free-text food descriptions into detailed nutritional profiles, NutriRAG can support diverse research and health monitoring applications, enhancing dietary tracking and nutritional insights in multiple technological environments.

To evaluate the clinical relevance of the NutriRAG’s capabilities to classify food groups, we analyzed eating habits in response to different types of dietary restriction. Our research noted distinct dietary pattern shifts in both TRE and CRE groups. The TRE group experienced substantial reductions in meal consumption, including breakfast, lunch, and late-night eating. In contrast, the CRE group showed marked decreases in savory snacks and high-added-sugar foods. Our results underscore the efficacy of TRE and CRE approaches in lowering energy intake [41]. Moreover, we observed that higher number of breakfast EOs are associated with lower insulin sensitivity as evidenced by lower HOMA scores. These findings highlight the importance of the timing and frequency of food consumption as key factors influencing metabolic health, particularly insulin resistance.

By leveraging daily food intake data from the app tracker, our analysis with NutriRAG between baseline and follow-up data demonstrated that both TRE and CRE groups decreased their daily meal occasions. This study provides further evidence supporting the efficacy of TRE in modifying dietary behaviors, emphasizing the potential of targeted eating schedules in enhancing metabolic health outcomes.

This study introduces a novel retrieval-based LLM framework designed for identifying food items and classifying food categories, specifically aimed at analyzing personal eating patterns. The study demonstrates the potential of retrieval-augmented LLMs in nutritional analysis. However, it also recognizes a limitation: the dependency on high-quality data for effective retrieval, which can be challenging in situations where relevant external data are limited or biased.

## CONCLUSION

NutriRAG represents a significant step forward in the application of AI in nutrition and health sciences. By effectively combining the advanced capabilities of LLMs with the contextually rich data retrieval, NutriRAG enhances the precision of identifying food items and classifying their categories while presenting an LLM-driven method to analyze complex dietary patterns in diverse populations. Future studies should focus on refine these models further, ensuring they are accessible and effective across different demographic and clinical settings.

## Data Availability

The data used in this study are not publicly available due to privacy and ethical restrictions, as they contain sensitive information from human participants. However, the NutriRAG model developed in this study is available upon reasonable request to the authors.

## CONTRIBUTORSHIP STATEMENT

HZ, LC, LH, RZ conceptualized and designed the study. HZ, LC, and LH curated the data and executed the experiments. HZ and LC drafted the initial manuscript, and all authors reviewed and finalized the manuscript.

